# Analysis of Race, Sex, and Language Proficiency Disparities in Documented Medical Decisions

**DOI:** 10.1101/2024.07.11.24310289

**Authors:** Hadi Amiri, Nidhi Vakil, Mohamed Elgaar, Jiali Cheng, Mitra Mohtarami, Adrian Wong, Mehrnaz Sadrolashrafi, Leo A. Celi

## Abstract

**Key Points:** 

**Question:** Are there disparities associated with race, sex, or language proficiency of patients in documented medical decisions within discharge summaries?

**Finding:** This study included expert annotation of 56,759 medical decisions across 451 discharge summaries reveals significant disparities associated with language proficiency of patients across different types of medical decisions in discharge summaries of specific disease groups.

**Meaning:** Disparities associated with sex and language proficiency of patients are present in the documentation of medical decisions, and addressing such disparities might promote equitable care and prevent computational models from learning and perpetuating such biases.

**Importance:** Detecting potential disparities in documented medical decisions is a crucial step toward achieving more equitable practices and care, informing healthcare policy making, and preventing computational models from learning and perpetuating such biases.

**Objective:** To identify disparities associated with race, sex and language proficiency of patients in the documentation of medical decisions.

**Design:** This cross-sectional study included 451 discharge summaries from MIMIC-III, with all medical decisions annotated by domain experts according to the 10 medical decision categories defined in the Decision Identification and Classification Taxonomy for Use in Medicine. Annotated discharge summaries were stratified by race, sex, language proficiency, diagnosis codes, type of ICU, patient status code, and patient comorbidities (quantified by Elixhauser Comorbidity Index) to account for potential confounding factors. Welch’s t-test with Bonferroni correction was used to identify significant disparities in the frequency of medical decisions.

**Setting:** The study used the MIMIC-III data set, which contains de-identified health data for patients admitted to the critical care units at the Beth Israel Deaconess Medical Center.

**Participants:** The population reflects the race, sex, and clinical conditions of patients in a data set developed by previous work for patient phenotyping.

**Main Outcomes and Measures:** The primary outcomes were different types of disparities associated with language proficiency of patients in documented medical decisions within discharge summaries, and the secondary outcome was the prevalence of medical decisions documented in discharge summaries. The data set will be made available at https://physionet.org/

**Results:** This study analyzed 56,759 medical decision text segments documented in 451 discharge summaries. Analysis across demographic groups revealed a higher documentation frequency for English proficient patients compared to non-English proficient patients in several categories, suggesting potential disparities in documentation or care. Specifically, English proficient patients consistently had more documented decisions in critical decision categories such as “Defining Problem” in conditions related to circulatory system and endocrine, nutritional and metabolic diseases. However, this study found no significant disparities in medical decision documentation based on sex or race.

**Conclusions and Relevance:** This study illustrates disparities in the documentation of medical decisions, with English proficient patients receiving more comprehensive documentation compared to non-English proficient patients. Conversely, no significant disparity was identified in terms of sex or race. These findings suggest a potential need for targeted interventions to improve the equity of medical documentation practices so that all patients receive the same level of detailed care documentation and prevent computational models from learning and perpetuating such biases.

## 1. Introduction

Disparities in medical decision making can impact patient outcomes and affect the quality and fairness of care.^[1]^ A discharge summary contains the key medical decisions for a patient’s hospital stay and serves as a method to communicate to other members of the health care team the reasons for hospitalization and patient’s subsequent hospital course to allow for improved transitions of care.^[2]^ The Decision Identification and Classification Taxonomy for Use in Medicine (DICTUM)^[3]^ categorizes medical decisions into 10 types and provides a comprehensive taxonomy of medical decisions.^[4—7]^ Despite extensive research on race and sex disparities in healthcare, less is known about the extent of these disparities in the documentation of electronic health records (EHR), particularly across other axes such as language proficiency of patients. Using DICTUM, this study provides a framework for categorizing medical decisions in discharge summaries and analyzing disparities associated with race, sex, and language proficiency across different medical decisions categories and patient groups. The goal of this study is to uncover patterns of disparities that might shed light on current practices and inform future interventions to improve equity in medical decision making and documentation. The implications of this research include informing healthcare policies for reducing disparities and improving overall healthcare delivery as well as mitigating relevant biases in computational models applied to healthcare data.

## 2. Method

This study included discharge summaries from MIMIC-III^[8]^ that were previously developed^[9]^ for patient phenotyping. Two domain experts independently read these discharge summaries and identified all text segments that contain medical decisions according to the 10 medical decision categories defined in DICTUM^[3]^ (see definitions in **Table 1**). A third annotator adjudicated any disagreements to ensure the accuracy of the annotations. All annotators were compensated. The inter-annotator agreement was measured by Cohen’s Kappa based on the token (word)-level agreement between the first two annotators.

**Table 1.**
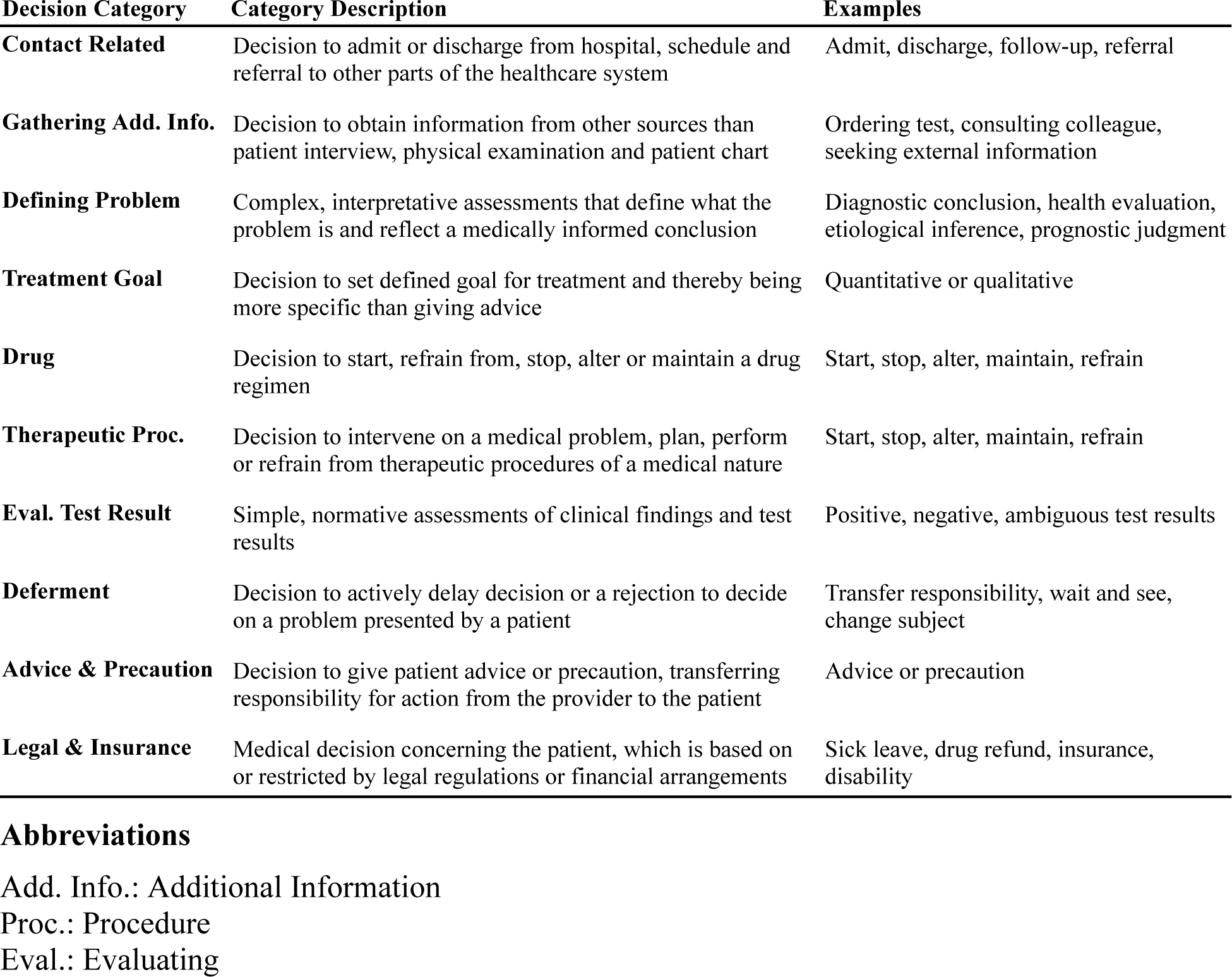
Medical Decision Categories in the Decision Identification and Classification Taxonomy for Use in Medicine (DICTUM).^[3]^.

The annotated discharge summaries were stratified by race, sex, language proficiency, diagnosis (ICD-9 codes), type of ICU, patient status code, and patient comorbidities (quantified by Elixhauser Comorbidity Index).^[10]^ This stratification was performed to reduce the chance that any observed disparities in documented medical decisions were due to variations attributable to these potential confounders. A Welch’s t-test, chosen for its robustness in handling unequal sample sizes and variances, was employed to identify significant differences in the frequency of specific medical decisions across these groups. Bonferroni correction was applied to adjust p-values (0.01 and 0.05 thresholds); this approach was chosen because it is more conservative and is particularly useful when the cost of a type I error (falsely declaring significance) is high.

## 3. Results

### 3.1. Annotation Results

The token (word)-level inter-annotator agreement between the first two annotators was substantial, Cohen’s Kappa of *k* = 0.74, indicating the clarity with which medical decisions can be categorized based on DICTUM. The annotated data set, summarized in **Table 2**, comprises 56,759 medical decision text segments from 451 discharge summaries, of which 4.4% (2,519 text segments) contained overlapping decisions (see an example in **Figure 1**). Decision categories such as “defining problem,” “drug,” “evaluation,” and “therapeutic procedure” accounted for the majority of decisions, showing substantial (Cohen’s Kappa *k*>0.61) to almost perfect (*k*=0.93 for drugs) token-level agreement between the first two annotators. In contrast, decision categories like “gathering additional information,” “treatment goal,” “deferment,” and “legal & insurance” were less common (less than 1% of the decisions), had poor inter-annotator agreement, and required frequent adjudication to ensure accuracy. Furthermore, text segment length varied significantly across decision categories. Decisions requiring detailed explanation, such as “advice” and “evaluation,” resulted in longer text segments, whereas procedural decisions like those in the “therapeutic procedure” category were more concise.

**Figure 1.**
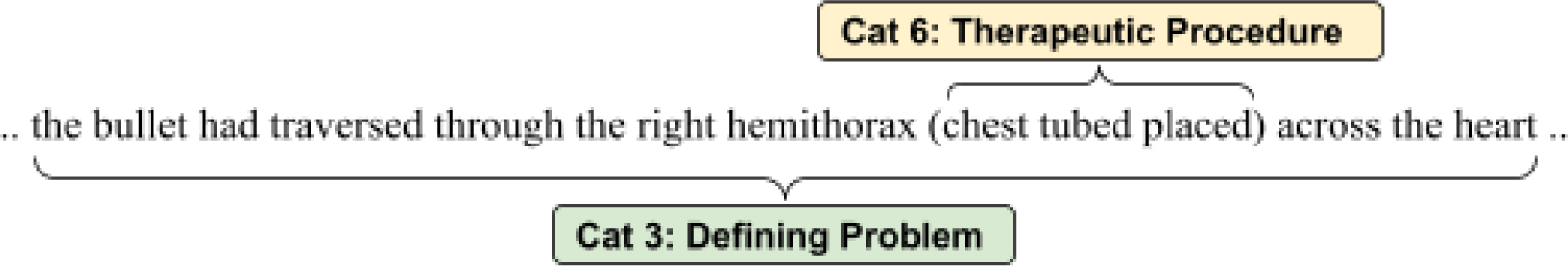
Example of overlapping medical decisions.

**Table 2.** Distribution of medical decisions and their corresponding annotation agreements in the data set. Decision Category (*Count*, *%*) indicates the count and percentage of medical decisions for each decision category. Agreement indicates token-level inter-annotator agreement measured based on Cohen’s Kappa between the first two annotators for each decision category. All disagreements were adjudicated to ensure the accuracy of the annotations.

| **Decision Category** | **(Count; %)** | **Agreement** | **Average Length of Decision Text (SD)** |
| --- | --- | --- | --- |
| **Defining Problem** | (22,289; 39) | 0.64 | 8.3 (8.7) |
| **Drug** | (14,569; 26) | 0.93 | 11.3 (12.3) |
| **Eval. Test Results** | (7,509; 13) | 0.68 | 15.4 (13.5) |
| **Therapeutic Proc.** | (6,958; 12) | 0.60 | 6.6 (7.1) |
| **Contact Related** | (2,872; 5) | 0.79 | 22.1 (25.7) |
| **Advice & Precaution** | (1,828; 3) | 0.77 | 24.7 (19.3) |
| **Gathering Add. Info.** | (454; 1) | 0.22 | 10.6 (9.5) |
| **Treatment Goal** | (169; 0) | 0.12 | 7.9 (4.8) |
| **Deferment** | (107; 0) | 0.23 | 11.1 (7.7) |
| **Legal & Insurance** | (4; 0) | 0 | 10.5 (6.3) |
| **Overall** | (56,759;100) | 0.74 | 8.4 (8.3) |

### 3.2. Disparity Analysis

**Table 3** summarizes the distribution of documented medical decisions across race, sex and language proficiency groups. The majority of discharge summaries were of male patients (n=259, 57.4%). The racial distribution was mostly White (77%), with fewer individuals identified as Black, Hispanic, Asian, or other race groups. Notably, no American Indian/Alaska Native individuals were represented. The majority of patients with known language proficiency were English speakers (85.2%), with a smaller group of non-English speakers (14.8%). The normalized average counts for sex and race groups showed comparable documentation frequency. However, the data shows a consistent trend of more medical decisions being documented for English speakers compared to non-English speakers across almost all categories.

**Table 3.** Distribution of medical decisions across target variables for disparity analysis. The values indicate the count of medical decisions of specific decision categories for each sex, race and language proficiency groups. (n) shows the number of patients in each category.

| Decision | Sex |  | Race |  |  |  |  |  |  | Lng. Prof. |  |
| --- | --- | --- | --- | --- | --- | --- | --- | --- | --- | --- | --- |
|  | Male<br>(n=259) | Female<br>(n=192) | White<br>(n=322) | AA<br>(n=42) | Hispanic<br>(n=23) | Asian<br>(n=12) | AI/AN<br>(n=0) | NH<br>(n=1) | Other<br>(n=18) | En<br>(n=260) | Non-En<br>(n=45) |
| **Defining Problem** | 12,950 | 9,405 | 16,069 | 2,132 | 1,174 | 511 | 0 | 29 | 973 | 14,337 | 2,465 |
| **Drug** | 8,604 | 6,087 | 10,437 | 1,385 | 795 | 348 | 0 | 18 | 715 | 9,656 | 1,612 |
| **Eval. Test Results** | 4,265 | 3,298 | 5,132 | 946 | 406 | 177 | 0 | 24 | 333 | 4,857 | 872 |
| **Therapeutic Proc.** | 4,040 | 3,009 | 5,004 | 710 | 359 | 190 | 0 | 10 | 325 | 4,426 | 756 |
| **Contact Related** | 1,636 | 1,266 | 2,039 | 264 | 176 | 75 | 0 | 8 | 120 | 1,795 | 320 |
| **Advice & Prec.** | 1,122 | 843 | 1,404 | 182 | 132 | 53 | 0 | 0 | 110 | 1,458 | 190 |
| **Gathering Info.** | 261 | 214 | 325 | 38 | 39 | 23 | 0 | 5 | 24 | 330 | 38 |
| **Treatment Goal** | 104 | 66 | 121 | 15 | 12 | 3 | 0 | 0 | 5 | 92 | 28 |
| **Deferment** | 71 | 44 | 83 | 10 | 7 | 0 | 0 | 0 | 3 | 71 | 14 |
| **Legal & Insurance** | 1 | 3 | 2 | 2 | 0 | 0 | 0 | 0 | 0 | 4 | 0 |

The analysis of medical decisions across various patient groups showed significant disparities based on language proficiency in the category of “Defining Problem” (**Table 4**). In defining problems within circulatory system diseases, and metabolic and immunity disorders, English proficient patients had substantially more decisions recorded (1,812 vs. 1,036 and 1,550 vs. 1,100 respectively), both with adjusted p-values of <0.01. This pattern was consistent across other conditions such as other metabolic and immunity disorders, where English proficient patients had significantly more decisions documented than their non-English counterparts (1,347 vs. 571, p<0.01). However, no significant disparities were found in medical decision documentation based on the sex or race variable (see Section 5, Limitations).

**Table 4.** Statistically significant disparities in documented medical decisions. “En” and “NonEn” indicate English proficient and Non-English patients respectively. “Gr” is the abbreviation for Group, and # indicates counts. For example, the first row shows two patient groups both diagnosed with conditions categorized under ICD codes 390—459 (diseases of circulatory system), type of ICU (CCU), Code Status (Full code), and Elixhauser Score (Low risk, lower 33%). Group 1 consists of 26 English proficient patients and Group 2 consists of 22 Non-English proficient patients. A total of 1,812 and 1,036 “Defining Problem” decisions were documented in the discharge summaries of these two respective groups. The p-values were adjusted using the Bonferroni correction based on thresholds set at 0.01 and 0.05. For all identified groups in this Table, the type of ICU is CCU, code status is Full Code, and Elixhauser score is low risk, lower 33%.

| Decision | ICD Code | Group1 | #Patients | #Decisions | Group2 | #Patients | #Decisions | Stats | p-value |
| --- | --- | --- | --- | --- | --- | --- | --- | --- | --- |
| Def. Problem | 390-459<sup>a</sup> | En | 26 | 1,812 | NonEn | 22 | 1,036 | 5.07 | 0.000007** |
| Def. Problem | 240-279<sup>b</sup> | En | 22 | 1,550 | NonEn | 22 | 1,100 | 4.43 | 0.000065** |
| Def. Problem | 270-279<sup>c</sup> | En | 19 | 1,347 | NonEn | 13 | 571 | 4.49 | 0.0001340* |

<sup>a</sup> Diseases of the Circulatory System
<sup>b</sup> Endocrine, Nutritional And Metabolic Diseases, And Immunity Disorders
<sup>c</sup> Other Metabolic Disorders And Immunity Disorders
\*\* $p < 0.01$
\* $p < 0.05$

## 4. Discussion

The high level of inter-annotator agreement observed in this study was inline with the agreement reported in the original DICTUM study^[3]^ and its follow up works.^[4]^ This consistency substantiates the reliability of the annotation process. It is important to note that token-level agreement underestimates the true extent of agreement among annotators. Differences in the inclusion/exclusion of less relevant information, such as stopwords, within a decision segment can result in what appears to be only partial agreement by different annotators, even when annotators largely agree in their interpretation of the text.

In terms of specific medical decision categories, the substantial inter-annotator agreement on “defining problem,” “drug,” “evaluation,” and “therapeutic procedure” likely stems from standardized terminology and more concrete definitions of these categories. In contrast, decision categories like “gathering additional information,” “treatment goal,” “deferment,” and “legal/insurance” seldom documented in discharge summaries and had low inter-annotator agreement. This could be attributed to the subjective nature or less specific language in such decision categories.

These findings suggest cases where English proficient patients are more likely to have a higher frequency of medical decisions documented, indicating potential broader disparities in healthcare documentation practices, communication or both across language groups. This study emphasizes the need for further investigation to ensure equitable medical documentation across different language groups.

## 5. Limitations

This study has several limitations, which are the subject of our future work:

The data set of 451 patients from MIMIC-III is relatively small and may limit the observation of statistical significance across sex and race variables. Extending this approach to the full MIMIC dataset could provide a more comprehensive understanding of disparities across these axes and across a broader range of disease categories; this is because high resolution data reveal more distinctive patterns.^[11]^ This extension would be possible using the approach we recently developed for automatic extraction of medical decisions from discharge summaries.^[12]^

MIMIC-III is limited to a single institution, which may not represent other settings, particularly those with different patient demographics or healthcare practices.

Discharge summaries provide a concise overview of a patient’s hospital stay, but they may not have a full coverage on all medical decisions made during a patient’s stay. Important interim decisions, particularly those not directly related to the discharge diagnosis or primary treatment, may be omitted. This incomplete representation may skew the analysis of disparities.

Despite measures to ensure consistency and substantial annotation agreement (k = 0.74), variations in how annotators interpret and categorize medical decisions could introduce inconsistencies. The presence of overlapping decisions within the text segments (as shown in **Figure 1**) makes the annotation process more complex. In addition, misclassification or inconsistent coding could affect the results of this study.

Finally, while non-medical factors such as socioeconomic status, education level, and cultural background should not affect medical decision-making, they often play a crucial role in health outcomes and are documented in clinical text;^[13]^ and impact patient engagement, communication effectiveness, and even the assumptions and biases held by healthcare providers.^[14]^ This study did not control for these factors, which could potentially introduce bias into the documented medical decisions.

## 6. Conclusions

This study analyzed disparities in the documentation of medical decisions within discharge summaries, associated with race, sex, and language proficiency. Analysis across demographic groups revealed that English proficient patients consistently had more documented decisions in critical decision categories such as “Defining Problem” in conditions related to circulatory system and endocrine, nutritional and metabolic diseases. However, this study found no significant disparities in medical decision documentation based on sex or race. Addressing potential disparities is essential for achieving equitable healthcare practices and care. The results inform policy making and have the potential to prevent computational models from learning and perpetuating such biases. Future research can investigate the underlying causes of these disparities and develop bias mitigation strategies.

## Data Availability

The data set will be made available at https://physionet.org/ and relevant tools at https://github.com/CLU-UML/MedDec

## Acknowledgment

The authors of this work report no conflicts of interest.

